# DUNE: a versatile neuroimaging encoder captures brain complexity across three major diseases: cancer, dementia and schizophrenia

**DOI:** 10.1101/2025.02.24.25322787

**Authors:** Thomas Barba, Bryce A. Bagley, Sandra Steyaert, Francisco Carrillo-Perez, Christoph Sadée, Michael Iv, Olivier Gevaert

**Author notes:** Corresponding authors: 1) Prof Olivier Gevaert, PhD Associate Professor of Medicine and Biomedical Data Science Stanford University 2) Dr Thomas Barba, MD, PhD Assistant Professor of Internal Medicine Department of Internal Medicine Lyon University Hospital, France.

## Abstract

Magnetic resonance images (MRI) of the brain exhibit high dimensionality that pose significant challenges for computational analysis. While models proposed for brain MRIs analyses yield encouraging results, the high complexity of neuroimaging data hinders generalizability and clinical application. We introduce DUNE, a neuroimaging-oriented encoder designed to extract deep-features from multisequence brain MRIs, thereby enabling their processing by basic machine learning algorithms. A UNet-based autoencoder was trained using 3,814 selected scans of morphologically normal (healthy volunteers) or abnormal (glioma patients) brains, to generate comprehensive low-dimensional representations of the full-sized images. To evaluate their quality, these embeddings were utilized to train machine learning models to predict a wide range of clinical variables. Embeddings were extracted for cohorts used for the model development (n=21,102 individuals), along with 3 additional independent cohorts (Alzheimer’s disease, schizophrenia and glioma cohorts, n=1,322 individuals), to evaluate the model’s generalization capabilities. The embeddings extracted from healthy volunteers’ scans could predict a broad spectrum of clinical parameters, including volumetry metrics, cardiovascular disease (AUROC=0.80) and alcohol consumption (AUROC=0.99), and more nuanced parameters such as the Alzheimer’s predisposing APOE4 allele (AUROC=0.67). Embeddings derived from the validation cohorts successfully predicted the diagnoses of Alzheimer’s dementia (AUROC=0.92) and schizophrenia (AUROC=0.64). Embeddings extracted from glioma scans successfully predicted survival (C-index=0.608) and IDH molecular status (AUROC=0.92), matching the performances of previous task-oriented models. DUNE efficiently represents clinically relevant patterns from full-size brain MRI scans across several disease areas, opening ways for innovative clinical applications in neurology.

**One Sentence Summary:** We propose a brain MRI-specialized encoder, which extracts versatile low-dimension embeddings from full-size scans.

## INTRODUCTION

The morphology of the human brain is determined by genetic and embryological factors specific to each individual, and is constantly remodeled throughout life in response to internal and environmental factors(1). A broad range of pathological processes shape brain morphology, either affecting the brain primarily (e.g., cancer and inflammatory and neurodegenerative diseases) or secondarily in systemic disorders (e.g., cardiovascular and systemic autoimmune diseases)(2–4). Therefore, the morphology of the brain recapitulates numerous formative events that occur during an individual’s lifetime, providing a snapshot of their health at a particular time. Advances in brain imaging over the last decades have enabled doctors to better characterize and treat a wide range of conditions. In this setting, magnetic resonance imaging (MRI) have established it as a fundamental tool in the diagnosis and surveillance of cancers, neurodegenerative disorders, and inflammatory diseases of the central nervous system(5,6). With the ongoing development of new imaging sequences, the quality of brain imaging continues to improve, achieving unprecedented precision in depicting brain morphology. However, while these increasingly complex representations of the brain are of undeniable interest in the management of brain diseases, their full potential has yet to be realized.

In the era of artificial intelligence, authors have proposed deep-learning models capable of leveraging these complex imaging data to predict clinical parameters such as detection of brain tumors(7,8), and diagnosis of neuropsychiatric and neurodegenerative diseases (9–11). However, these models have been limited in their clinical translation due to several reasons. First, their training requires large datasets and extensive computational resources because of the considerable number of internal parameters that need to be trained. Second, their monothematic nature implicates that they learn to perform only one task at a time (e.g. diagnosis prediction or tumor segmentation). Third, these models are usually trained on a few specific datasets, which affects their performance on externally acquired images, a problem known as overfitting and lack of generalization(12).

General-purpose feature extractors could offer promising solutions to tackle the complexities of processing high-dimensional data. Such models can generate versatile representations that capture essential information from the input data, while reducing computational demands for downstream applications (*13*) (*14*). We thus sought to develop a general-purpose feature extractor, aimed at deriving non-invasive biomarkers from neuroimaging data that could be readily utilized by simple machine learning models, thereby facilitating their deployment in clinical settings even with limited data availability.

In this study, we introduce *DUNE* (**D**eep feature extraction by **U**Net-based **N**euroimaging-oriented auto**E**ncoder), a neuroimaging-oriented deep-learning model capable of deriving deep features from multisequence MRI scans. To extract a low-dimensional and versatile embeddings from full-sized brain scans, we used a UNet-based autoencoder, trained in an unsupervised manner to encode and reconstruct MRI data (15–18). To ensure its robustness, the model was trained on 3,814 MRI scans of morphologically normal and abnormal brains issued from three datasets. The quality of the extracted embeddings was evaluated by employing them to infer numerous clinical parameters using simple machine learning models. Different autoencoder architectures were compared in terms of reconstruction capabilities and quality of embeddings. Based on comprehensive evaluations across multiple clinical applications, we identified a UNet architecture without skip connections (U-AE) as the optimal model for DUNE. Surprisingly, this architecture, while being the least effective in reconstructing MRI data, produced the most clinically relevant embeddings, underscoring the importance of using appropriate evaluation metrics commensurate with their intended use. To demonstrate its generalizability, *DUNE* was used to extract brain MRI embeddings of 1,322 individuals from three independent cohorts representing three disease areas. These embeddings were used to successfully predict a broad spectrum of clinical parameters, including cognitive dysfunction, psychiatric disorders, genetic traits (APOE4 status), glioma molecular features (IDH status) and patient survival, with results matching or outperforming previously published models directly processing the raw imaging data. Finally, we demonstrate that augmenting the MRI data with synthetic images can enhance the quality of the produced embeddings, resulting in improved performances in downstream clinical inference tasks. In summary, *DUNE* provides efficient general-purpose low-dimensional embeddings of multi-modal brain MRI data independent of the downstream use case.

## RESULTS

### Workflow for development and evaluation of DUNE

We introduce *DUNE*, a framework designed to perform **D**eep feature extraction by **U**Net-based **N**euroimaging-oriented auto***E***ncoder. An overview of the workflow for its development and evaluation is presented in Fig 1a. First, we trained the model to extract low-dimensional embeddings from full-sized brain MRI scans. Subsequently, the quality of these embeddings was assessed by using them as inputs to machine learning models tasked with clinical inference and evaluating the resultant predictive performance on these downstream tasks.

**Figure 1.**
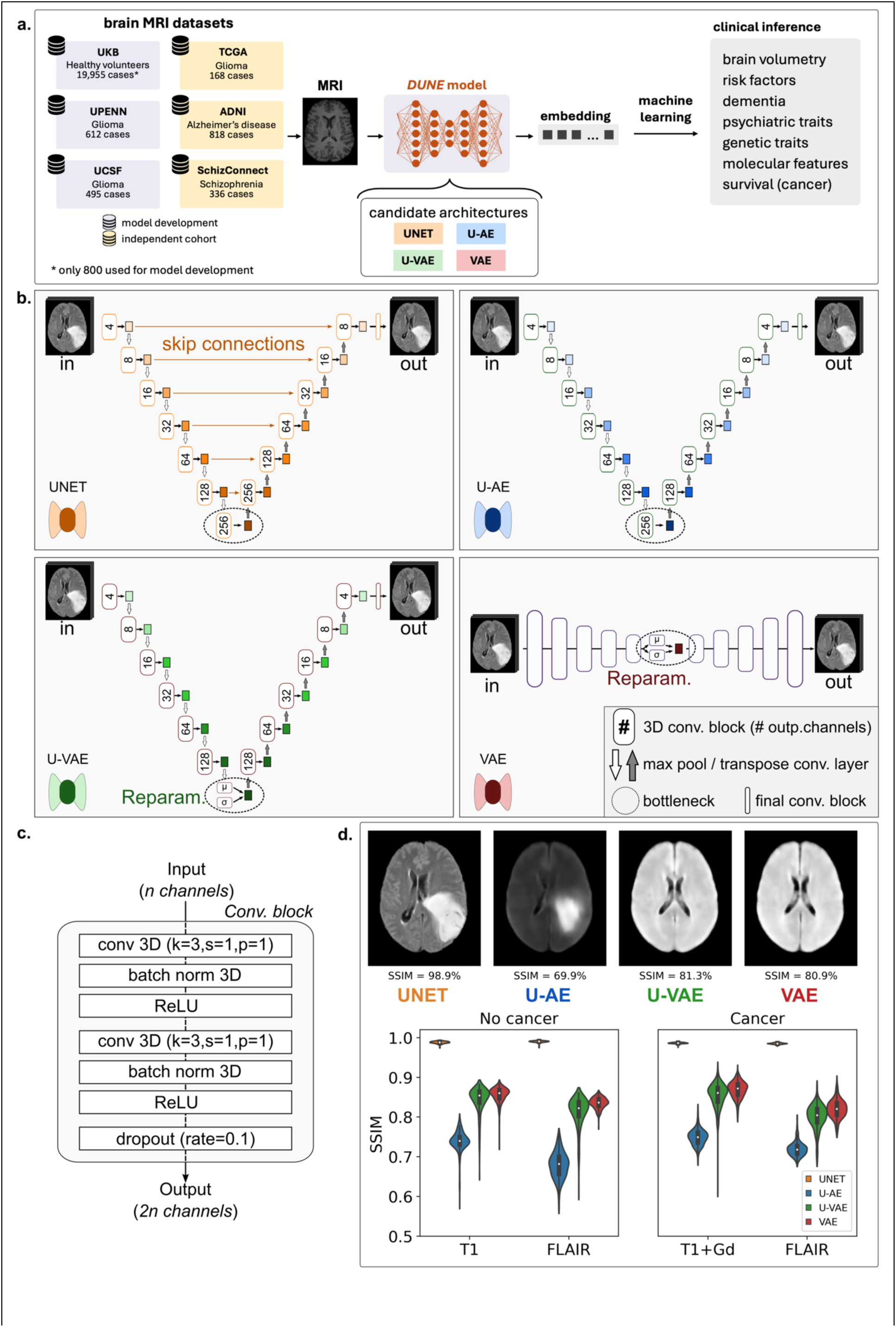
Project overview and model development. a) Workflow for the development and evaluation of the *DUNE* model. The development of the model involved three distinct datasets: UKB, UPENN, and UCSF. The *DUNE* model trained to encode and reconstruct full-sized brain MRI scans, thereby generating low-dimensional embeddings that capture the salient information from the original images. Subsequently, embeddings were generated for scans from the development datasets as well as three independent cohorts (TCGA, ADNI, SchizConnect). These compact representations were then utilized as inputs to simple machine learning models for the inference of clinical variables, allowing for an assessment of the embeddings’ quality and their ability to facilitate downstream clinical applications. b) We benchmarked four distinct candidate architectures for *DUNE*: UNET, U-AE, U-VAE, and VAE. Three of the models (UNET, U-AE, and U-VAE) are based on the U-Net architecture, while VAE is a variational autoencoder model adopted from a previous study(*18*). Specifically, UNET is the foundational U-Net model, U-AE is a variant without skip connections, and U-VAE is a variational version of U-AE that generates embeddings through a reparameterization procedure. c) The models are composed of sequential convolutional blocks and max pooling layers that reduce the dimension of brain MRI to generate the embeddings. d) The base UNET model demonstrated the highest similarity (SSIM) between the output and original images. While the U-AE generated less precise images, it appeared to better identify cancer characteristics compared to variational autoencoders, which failed to reconstruct the tumor.

Several datasets served for development and validation of *DUNE* (Fig 1a, Supplemental Table S1). The model development was made using brain MRI scans from the UKB, UPENN and UCSF datasets. The UKB (UK Biobank) dataset provides extensive clinical and radiological data for a large cohort of approximately 500,000 healthy individuals. Brain MRI (T1, FLAIR), and clinical data of 19,955 individuals were downloaded and preprocessed as described in the Methods section. To prevent overfitting the model to normal healthy brains, only 800 randomly selected cases were finally kept for the model development (n=1,600 T1 and FLAIR scans). In addition to these healthy brains were included scans (T1+gadolinium [T1Gd] and FLAIR) from the glioma cohorts UPENN (UPenn-GBM, n=1,224 scans) and UCSF (UCSF-PDGM, n=990 scans), to make the model equally capable of extracting features from morphologically abnormal brains. As a result, 3,814 MR 3D-images (T1, T1Gd, FLAIR) were used for training (80%) and validation (20%) purposes.

Four candidate architectures were systematically benchmarked to identify the optimal feature extractor for brain MRI. Guided by previous research (*19*), we focused our investigation on unsupervised autoencoders, given their potential for learning efficient and versatile embeddings. The architectures of these competing models are detailed in the Methods section and depicted in Fig 1b-c. We utilized autoencoders based on the UNet architecture, given their previous successes in medical images processing, (*20,21*). The first model was a “vanilla” 3D UNet autoencoder (UNET). Second, we reasoned that the UNet skip connections, while helpful in supervised segmentation tasks(*20*), could be detrimental to the embedding quality in our application. We therefore derived a version of the UNET model without skip connections (U-AE). Third, given published data suggesting that variational autoencoders can be useful for MR feature extraction(*22,23*), we designed a variational version of the U-AE model (U-VAE), by including a reparameterization layer in the bottleneck section. Finally, we used a fully connected variational autoencoder (VAE) which successfully extracted features from CT images of lung lesions in a previous study(*18*).

All four models trained to encode and reconstruct single-sequence 3D brain MRI scans (Fig 1d). Overall, the UNET model outperformed the other autoencoders in reconstructing brain MR with the best performance (SSIM=98.7 ±4.2%), all sequences (T1, T1Gd, FLAIR) considered, for images with cancer and non-cancer disorders (Fig 1d). Variational autoencoders (U-VAE and VAE) showed similar results for image reconstruction (82.6 ±4.5% and 84.4 ±2.8% respectively) but notably failed to reconstruct brain tumors (Fig 1d). The classical U-AE autoencoder demonstrated the worst reconstruction abilities (SSIM=72.2 ±4.2%).

### Canonical Correlation Analyses (CCA) reveal associations between MRI embeddings and clinical phenotypes

To identify the optimal architecture for DUNE among our four candidates, we conducted a comprehensive evaluation based on the quality of the embeddings they produce. As a first assessment criterion, we measured how well these embeddings correlated with clinical variables (Fig 2). We used all four candidate models to encode the images from the UKB dataset, with radiomics serving as a control given its established role as a state-of-the-art method for image feature extraction(*24*). For each patient (n=19,955), we generated global brain MR embeddings by concatenating features extracted from both T1 and FLAIR sequences.

**Figure 2.**
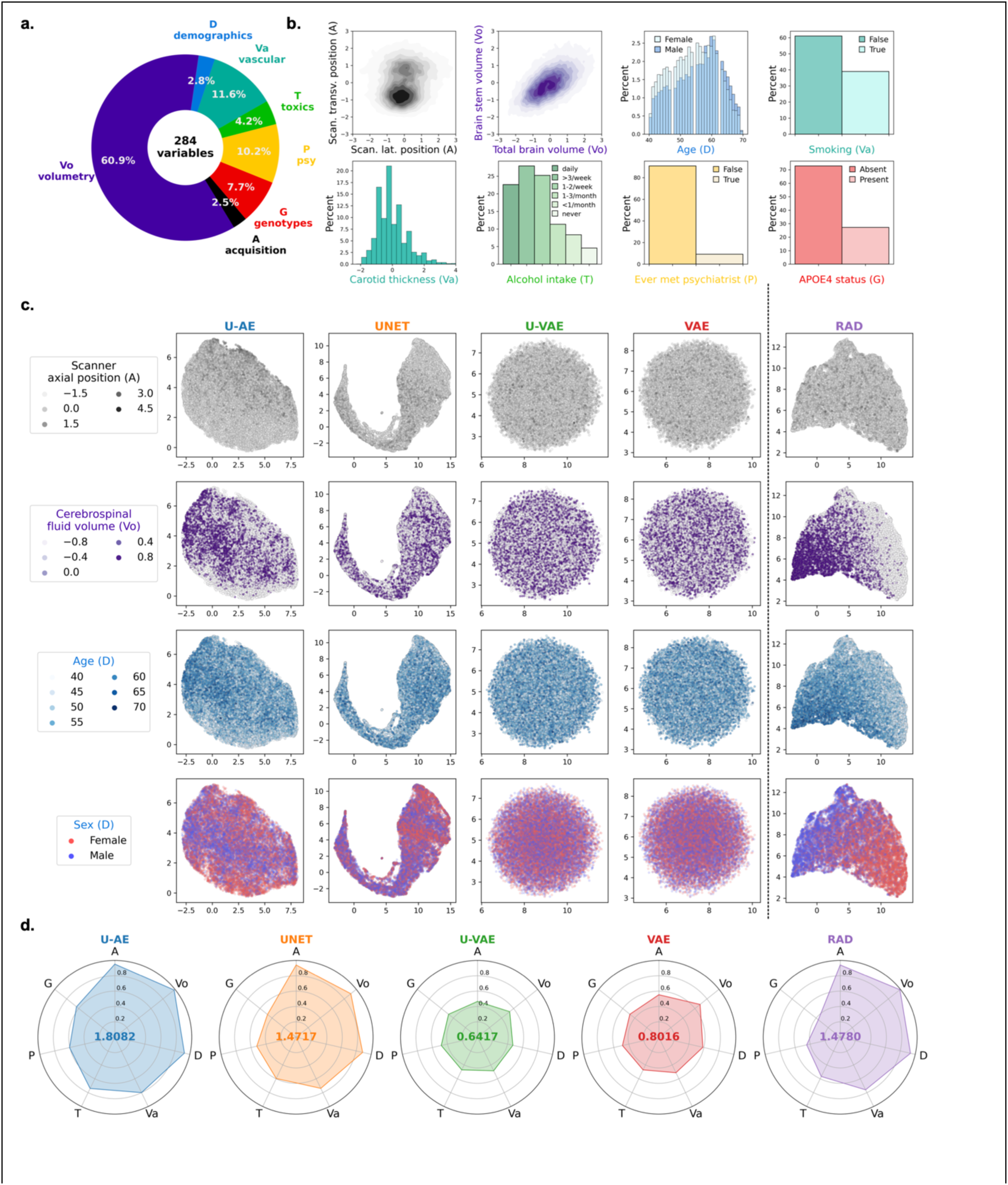
MRI embeddings correlate with clinical phenotypes in healthy individuals. a) 284 clinical parameters collected in the UKB dataset were divided into 7 categories (Vo: volumetry, A: acquisition protocol, G: genotypes, P: psychiatric traits, T: toxics, Va: vascular conditions, D: demographics). b) Distribution of illustrative variables from the different categories. c) UMAP reduction was performed on embeddings extracted from brain MR images by the 4 models and on radiomics features as control. Radiomics, UNET and U-AE generated embeddings successfully capture variables of acquisition, volumetry and demographic categories, while those issued by variational autoencoders do not. d) Canonical correlation analysis (CCA) between embeddings and clinical variables from each category. The radar plots display the correlation (R^2^ metric) between the first canonical variate of each category and that of each embedding, showing that U-AE embeddings correlate best with UKB clinical parameters.

We analyzed these embeddings against 284 clinical variables hypothesized to be associated with brain morphology. These variables were categorized into seven distinct groups: acquisition parameters (A), brain volumetry (Vo), demographics (D), vascular risk factors (Va), toxics (T), psychiatric (P) and genetic (G) traits (Fig 2a and b). The variables showed diverse statistical distributions, with some quantitative variables such as volumetric measurements, age, and carotid thickness following normal distributions (Fig 2b), while certain qualitative variables, notably APOE4 status, exhibited significant class imbalance. Our correlation analyses revealed strong associations within each category but weak correlations between categories (Supplemental Figure S2), leading us to analyze each group independently.

UMAP analyses of the embeddings revealed distinct patterns (Fig 2c). The variational autoencoders (U-VAE and VAE) produced normally distributed embeddings, as expected given their underlying statistical constraints. Embeddings from radiomics, U-AE, and to a lesser extent UNET, showed clear relationships with key clinical parameters, evidenced by distinct color patterns corresponding to acquisition parameters (scanner table position), volumetric measurements (cerebrospinal fluid volume), and demographic features (age and sex). In contrast, the embeddings from variational autoencoders (VAEs) showed no discernible patterns in relation to these clinical variables.

To assess the overall relationship between embeddings and clinical variables, we performed Canonical Correlation Analysis (CCA) on each clinical variable group. Figure 2d displays the determination coefficients (R² scores) between the first canonical variates of the embeddings and that of each subgroup of clinical variables. The total area covered by these coefficients in the radar plot summarizes the overall correlation strength. Using this metric, U-AE embeddings demonstrated the strongest correlation with clinical variables (area=1.81), outperforming the radiomics features (area=1.48), which ranked second.

### MRI embeddings accurately predict clinical phenotypes of healthy individuals

As a second evaluation criterion, we assessed how well each type of embedding could serve as input features for clinical prediction tasks. To do this, we trained supervised machine learning models to predict each of the 284 clinical variables from the UKB dataset. For each variable, models were trained on embeddings from each architecture (UNET, U-AE, U-VAE and VAE) or radiomics (RAD), used as control. The performance of clinical prediction was measured by the R^2^ score (quantitative variables) or the weighted F1-score (categorical variables). The results are displayed in Fig 3a (overall performance, and per variable subgroups).

**Figure 3.**
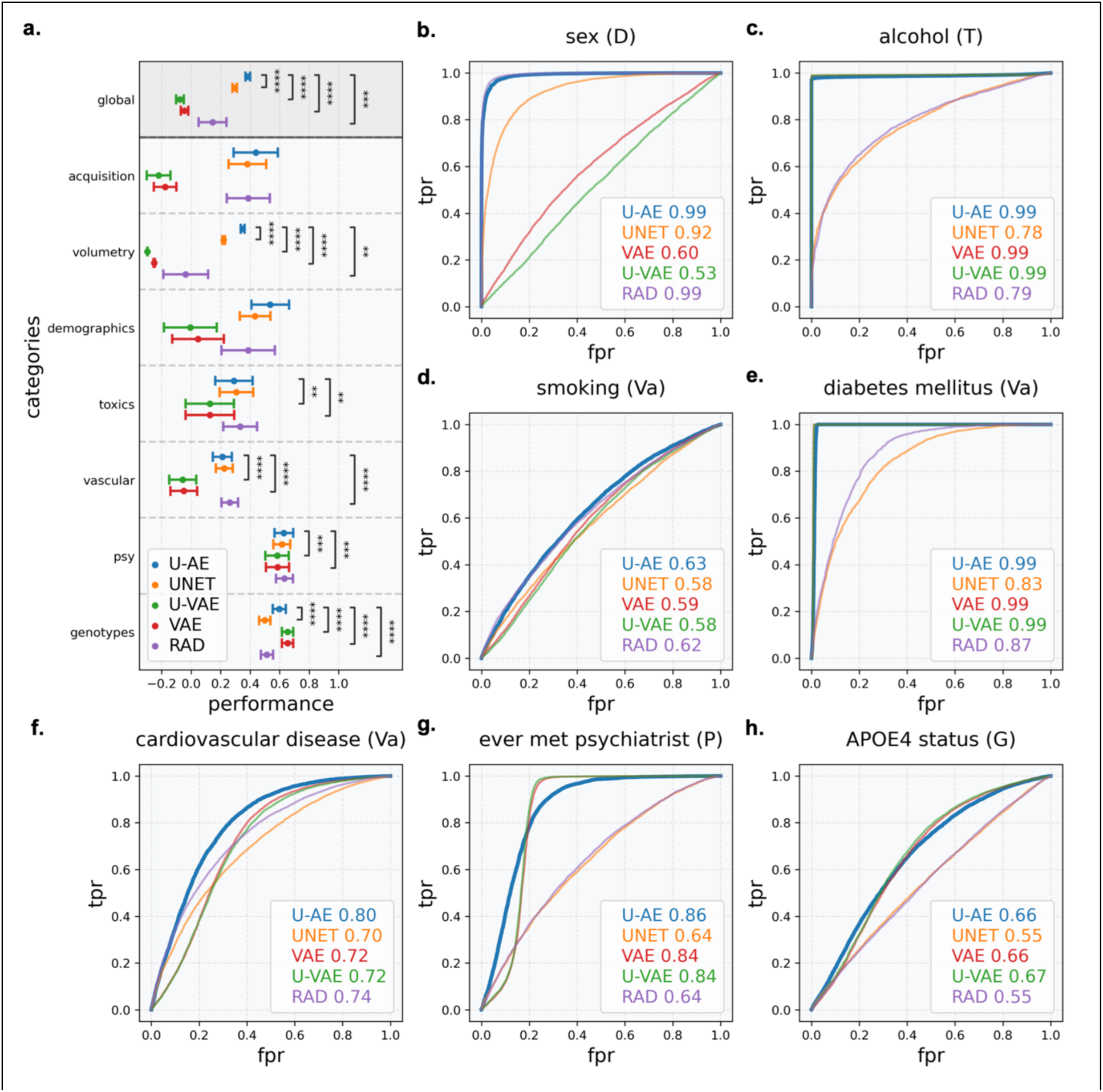
Clinical inference using MRI embeddings of healthy brains. a) Performance of machine learning models trained to predict clinical variables using model-generated embeddings of the UKB brain MRIs (T1 and FLAIR sequences, n=19,955 cases, 5-fold cross validation). Points and error bars show means and standard errors of prediction scores (R^2^ or F1-scores) across variables in each category. Overall, U-AE embeddings yielded the best predictions, except for toxics, genotypes and vascular conditions that were best predicted by radiomics and variational autoencoders respectively b) to h) Individual performance of clinical inference based on either autoencoder embeddings or radiomics.

Overall, the predictions were best with U-AE embeddings (0.384±0.27%), which significantly outperformed radiomics (0.144±1.58, p<0.001, Fig 3a). The embeddings encoded by other models (UNET, U-VAE, VAE) resulted in weaker predictions. The performances greatly varied between the different subgroups of clinical variables but showed small variation across variables within each category (Fig 3a), which tended to be highly intercorrelated (Supplemental Figure S2). Figure 3b to 3h thus display the performance of model predictions for illustrative variables from each category. U-AE embeddings were the most relevant in predicting most clinical variables. Embeddings encoded by variational autoencoders (U-VAE and VAE) were outperformed in all categories, except genetics.

### MRI embeddings capture cancer-specific features including molecular features and predict patient survival

Having validated our models on healthy brains, we next investigated their generalization capabilities to pathological cases. Specifically, we first focused on glioma patients, whose brain MRIs exhibit significant morphological abnormalities due to tumor presence. For this analysis, we used two categories of datasets: UCSF (n=495) and UPENN (n=612), which were part of the autoencoder training set, along with the previously unseen TCGA-GBM and TCGA-LGG datasets (n=168 combined). Once more, we assessed the quality of the embeddings in oncological context, by developing survival prediction models using them as input. For evaluation, we stratified the patients into two groups based on their predicted mortality risk (above or below median) and compared their survival curves. The prediction quality was measured by two complementary metrics: the Brier score, which measures prediction accuracy (optimal when close to 0), and the concordance index (C-index), which measures ranking accuracy (optimal when close to 1).

Models using U-AE encoded embeddings demonstrated superior performance in discriminating between low and high-risk patients. This superiority was consistent across both internal validation datasets, with C-index=0.587 and Brier score=0.184 for UPENN, and C-index=0.686 and Brier score=0.204 for UCSF, outperforming both radiomics features and alternative encodings from UNET and VAE architectures (Fig 4a and b). The external validation on the TCGA dataset confirmed these findings, with U-AE embeddings achieving better survival predictions (C-index=0.608) compared to radiomics (C-index=0.600) and other autoencoder-based approaches (Fig 4c).

**Figure 4.**
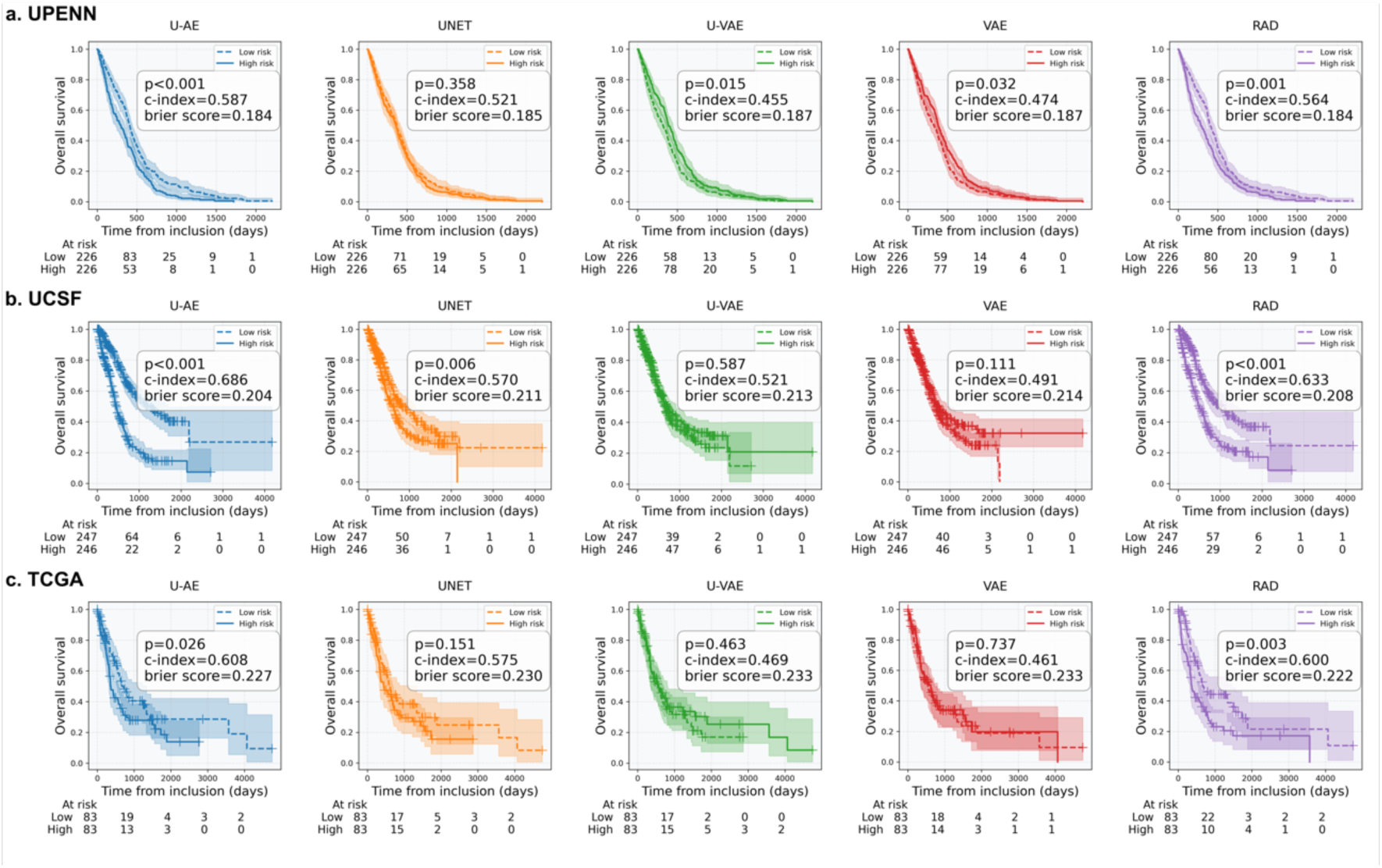
MRI embeddings allow for survival inference in individuals with glioma. Performance of random forest survival models predicting patient global survival based on MR embeddings/radiomics (T1Gd and FLAIR sequences) of 3 glioma datasets (5-fold cross validation). The overall survival of two groups, defined by risk scores of death below (low risk) or above (high risk) median, was compared (concordance index and log-rank test), for the UPENN (a) and UCSF (b) datasets (which were used as train sets for the autoencoders), and for the external TCGA (c) dataset. Predictions using the U-AE embeddings had the highest concordance index for all three datasets.

Beyond survival prediction, we evaluated the embeddings’ ability to predict key molecular characteristics of gliomas that guide therapeutic decisions. These included IDH1 mutation status and MGMT promoter methylation status, which are established markers of response to chemoradiation and temozolomide-based chemotherapies (Fig 5a and b). The U-AE embeddings consistently demonstrated superior predictive performance, achieving high accuracy in detecting IDH1 mutations (AUROC=0.99 in UCSF and AUROC=0.92 in TCGA), MGMT promoter methylation status (AUROC=0.77), and tumor grade (AUROC=0.94). Notably, even in these morphologically complex cases, the embeddings maintained their ability to predict general anatomical features such as patient sex (data not shown).

**Figure 5.**
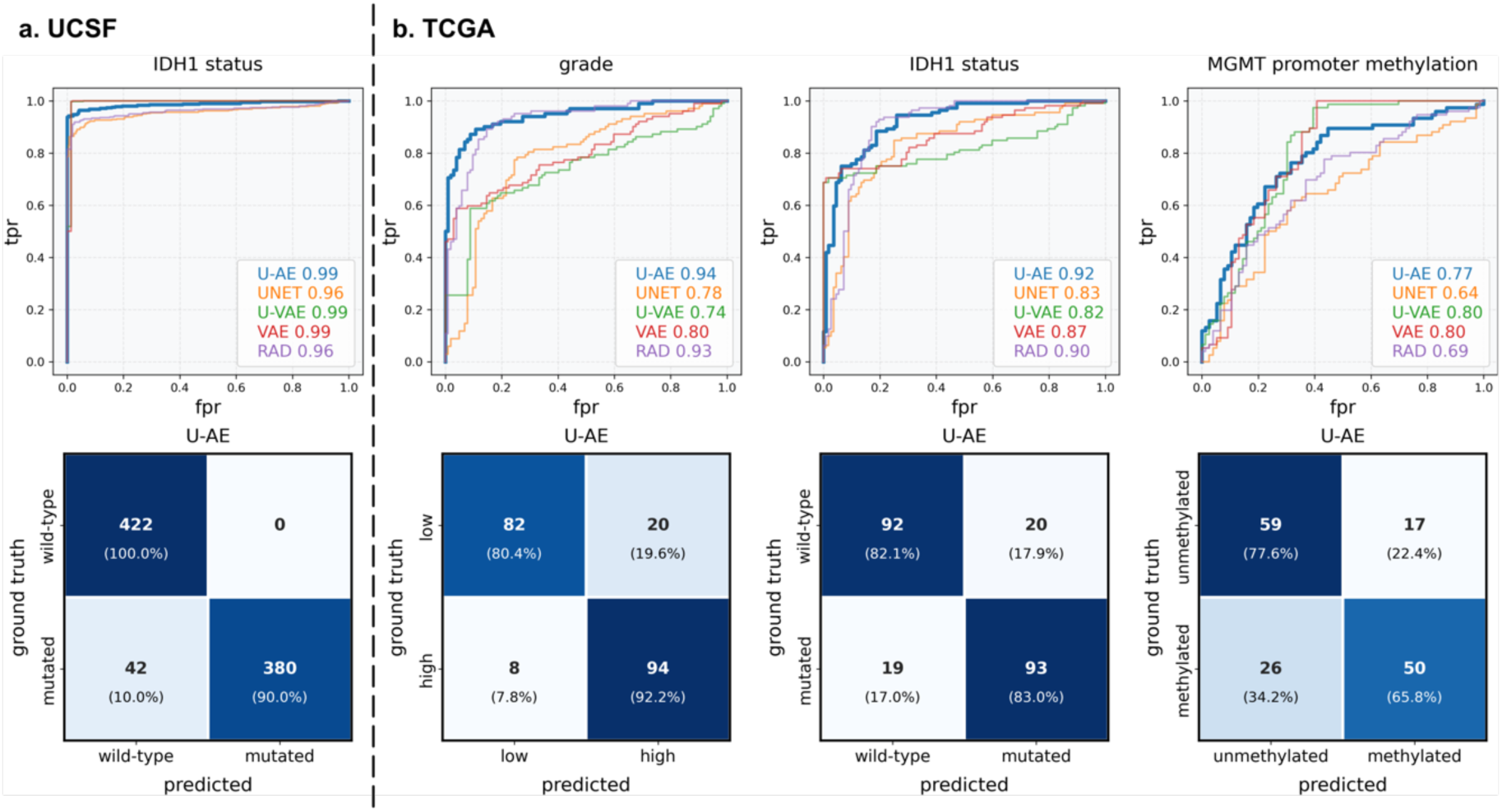
MRI embeddings enable inference of molecular features in patients with glioma. Performance of models predicting molecular features using MR embeddings/radiomics (T1Gd and FLAIR sequences) of the UCSF (a, IDH1 status) and TCGA datasets (b, cancer grade, IDH1 status, MGMT promoter methylation status).

### MRI embeddings predict diagnosis of neurodegenerative and psychiatric disorders and can be improved by synthetic data

We continued the benchmarking of the different models with two additional neurological conditions: neurodegenerative disease and schizophrenia. For this analysis, we leveraged data from the ADNI database for Alzheimer’s disease and the SchizConnect database (comprising COBRE and MCIC datasets). Unlike our previous analyses, these datasets contained only T1-weighted images, allowing us to test our models’ performance with limited imaging sequences.

In the Alzheimer’s disease cohort, we first evaluated the models’ ability to predict cognitive impairment severity. The U-AE embeddings demonstrated robust performance with an AUROC of 0.92 (Fig 6a). We then evaluated the prediction of genetic markers known to influence Alzheimer’s disease progression: the APOE4 allele status and the TOMM40 poly-T polymorphism length (dichotomized at the cohort median of 33). Interestingly, for these genetic predictions, the variational autoencoders performed the best. The U-VAE and VAE embeddings achieved perfect prediction for the APOE4 allele (AUROC=1) and moderately high accuracy for TOMM40 polymorphism length (AUROC=0.67 and 0.71, respectively), compared to the U-AE embeddings (AUROC=0.98 for APOE4 and AUROC=0.68 for TOMM40) (Fig 6a).

**Figure 6.**
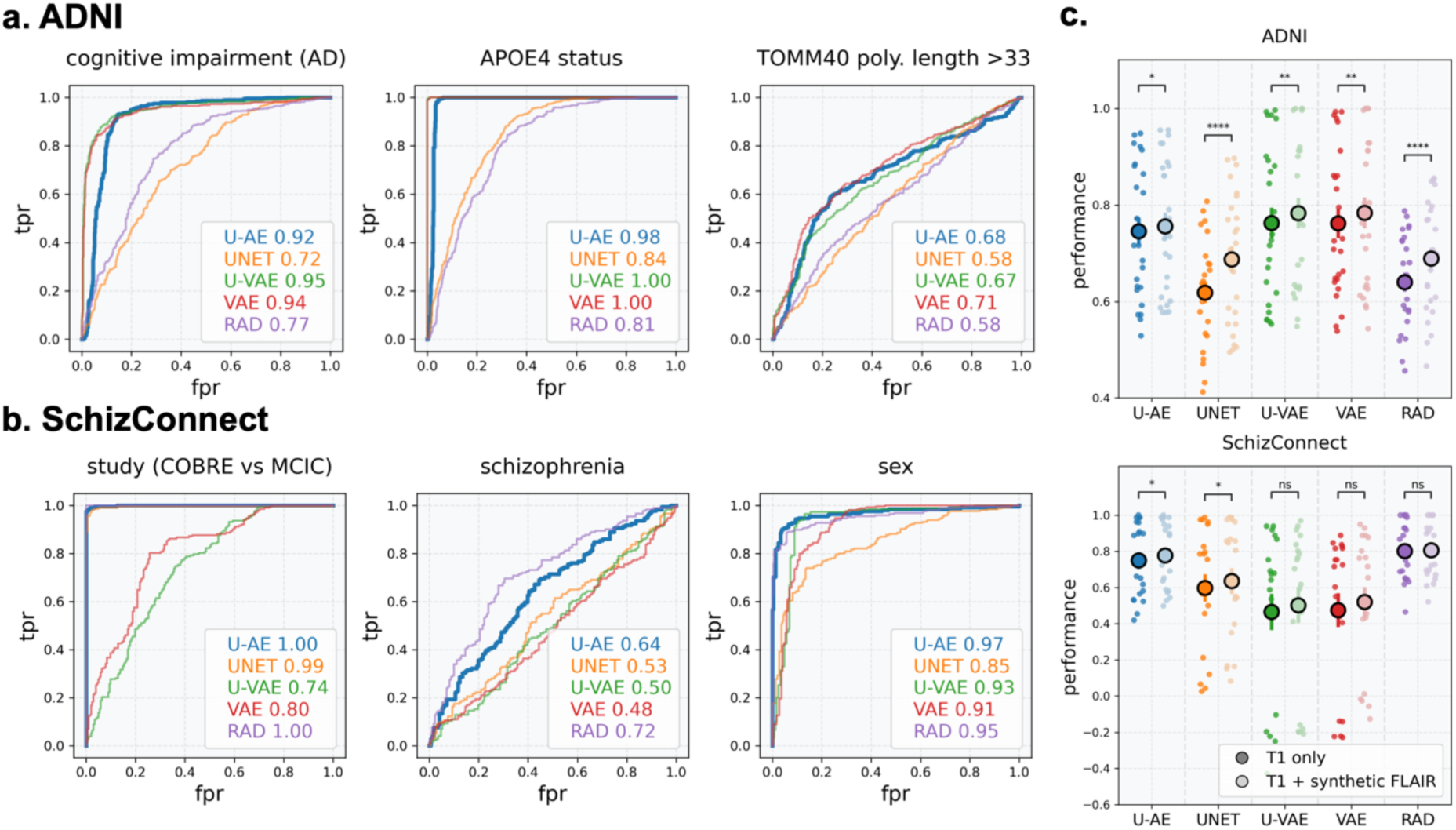
Clinical inference in Alzheimer’s disease and schizophrenia. a) Performance of models predicting clinical features (cognitive impairment [MMSE>28]) and genotypes (APOE4 status and length of the TOMM40 poly-T polymorphism [>33]) using MR embeddings/radiomics of the ADNI dataset (T1 sequences). b) Performance of models predicting parameters and clinical variables (cohort of origin [COBRE vs MCIC], schizophrenia vs healthy patients, sex and age) using MR embeddings/radiomics of the SchizConnect dataset (T1 sequences). c) Performance of models predicting clinical parameters from ADNI (top) and SchizConnect (bottom) datasets based on MR embeddings/radiomics of either T1 only images (dark dots) or both T1 + synthetic FLAIR images (light dots).

In the schizophrenia cohort, we assessed the models’ performance across four distinct prediction tasks: identification of study origin (COBRE vs MCIC, which reflected differences in MR acquisition protocols), clinical diagnosis (schizophrenia vs healthy volunteers), and demographic characteristics (sex and age). The U-AE embeddings maintained their superior performance, perfectly discriminating between studies of origin (AUROC=1) and accurately predicting patient sex (AUROC=0.97). For schizophrenia diagnosis, both U-AE embeddings and radiomics features achieved meaningful predictive performance (AUROC=0.64 and 0.72 respectively) (Fig 6b).

To address the limitation of single-sequence availability in the ADNI and SchizConnect datasets, we developed an enhancement strategy using synthetic data generation. We implemented a dedicated UNet-based model trained on the UK Biobank dataset (n=39,910 sequences, with an 80/20 train/validation split) to perform bidirectional synthesis between T1 and FLAIR sequences (Supplemental Figure S3). This approach proved highly effective, with synthetic FLAIR images showing strong concordance with real FLAIR images in the validation set (similarity index = 0.90 ± 0.04). We then applied this synthetic data enhancement to both the SchizConnect and ADNI datasets, generating FLAIR sequences from their available T1 images. This strategy enabled us to compare the predictive performance between embeddings derived from T1 images alone and those generated from the combination of T1 and synthetic FLAIR images, using our established clinical parameters.

The integration of synthetic FLAIR images yielded consistent improvements in prediction performance across all analyses. In the ADNI dataset, embeddings combining T1 and synthetic FLAIR sequences demonstrated enhanced predictive capability compared to those derived from T1 sequences alone (0.756±0.14 vs 74.5±0.14 with U-AE embeddings; Fig 6c top). We observed similar improvements in the SchizConnect dataset, where the addition of synthetic FLAIR sequences enhanced the predictive performance of the embeddings (0.777±18.9% vs 0.750±0.21 with U-AE embeddings; Fig 6c bottom). Remarkably, this enhancement in performance was consistent across all autoencoder architectures, suggesting that the benefit of incorporating synthetic FLAIR sequences extends beyond any specific model implementation.

## DISCUSSION

We introduce *DUNE*, a general-purpose feature extractor developed through systematic benchmarking of four autoencoder architectures. By evaluating these relatively simple architectures on diverse prediction tasks, we identified that a skip-connection-free UNet autoencoder (U-AE) consistently generates the most informative embeddings from brain MRIs. These embeddings demonstrate remarkable robustness across diverse patient cohorts – from healthy volunteers to those with gliomas, neurodegenerative diseases, and psychiatric disorders – capturing both readily apparent features, such as global brain morphology and demographics, and more subtle characteristics including genetic traits and molecular markers.

The versatility of DUNE’s embeddings was demonstrated across multiple levels of clinical prediction. When used with simple machine learning models, they enabled accurate prediction of basic parameters such as brain volumetry and demographic features (age, sex), while simultaneously capturing complex molecular and genetic signatures. In neuro-oncology, they achieved robust prediction of IDH1 mutation status, matching the performance of sophisticated deep learning models that process whole MR images directly (25–27). In neurodegenerative diseases, they successfully identified genetic traits such as APOE4 allele status, and in psychiatric disorders, they effectively detected subtle brain alterations associated with disease status.

Among the candidate architectures we evaluated, UNet-based autoencoders were initially selected for their relative simplicity and proven success in medical image processing (*28*). We hypothesized that such models could generate embeddings that faithfully capture the fundamental features of brain MRI structures. Unlike conventional applications, we employed unsupervised training through image reconstruction to optimize embedding versatility. Our systematic evaluation revealed that while the standard UNet architecture excelled at image reconstruction, its skip connections, which allow information to bypass the bottleneck layer, resulted in suboptimal embeddings. The removal of these skip connections in our U-AE model forced all information through the bottleneck, producing more informative embeddings despite lower reconstruction quality. We also explored variational autoencoders based on previous work (19,29–31), but these were outperformed by traditional architectures and even radiomics features, despite careful tuning of the Kullback-Leibler divergence term in the loss function.

Recent advances in computer vision have highlighted vision transformers as powerful tools for image analysis (*32,33*). Their self-attention mechanisms enable sophisticated spatial reasoning by weighing the importance of different image regions, which could theoretically benefit brain MRI analysis (*34,35*). However, several considerations led us to favor autoencoders for this application. First, while vision transformers excel at classification tasks, they are not inherently designed for dimensionality reduction and feature extraction, which was our primary objective. Second, the traditional autoencoder architecture provides an interpretable bottleneck structure that naturally aligns with our goal of generating compact, meaningful embeddings. Third, vision transformers typically require extensive pre-training on large-scale datasets to achieve optimal performance, making them less suitable for specialized medical imaging applications where data availability is limited. Finally, autoencoders offer better computational efficiency and have demonstrated superior adaptability to smaller, domain-specific datasets (*36*). This latter point was particularly crucial for our application, as it enabled us to train effectively on our relatively modest collection of brain MRI scans while maintaining model stability and generalization capability.

A major challenge in developing MRI analysis models is the heterogeneous nature of image acquisition across different centers. This heterogeneity makes image normalization particularly challenging and often leads to models that overfit to their training dataset and generalize poorly to external data. We addressed this challenge through two key strategies. First, we implemented a comprehensive image standardization pipeline, enabling our models to effectively learn from multiple datasets while maintaining robustness to external data. Second, and more importantly, we identified that maintaining a balanced representation of different brain morphologies in the training data was crucial. Initially, training with the complete UKB cohort (≈20,000 cases) alongside the glioma datasets led to a bias towards normal brain morphology, resulting in poor-quality embeddings for tumor cases. We resolved this issue by downsampling the UKB dataset to match the size of the glioma datasets, which improved the quality of tumor embeddings while preserving the model’s performance on normal brain images (data not shown).

The processing of multiple MRI sequences represented another key optimization challenge in our model development. Initially, we explored a multi-channel approach where different MR sequences were concatenated and processed simultaneously, similar to how RGB channels are handled in natural image processing. However, our systematic evaluation revealed superior performance with an alternative strategy: processing each MR sequence independently through separate encoding paths and subsequently concatenating their extracted features. This sequential approach not only improved the quality of the extracted features but also provided greater operational flexibility, allowing the model to handle incomplete imaging datasets where certain sequences might be unavailable. Such flexibility is particularly valuable in clinical settings, where the availability of multiple MRI sequences cannot always be guaranteed.

Literature is scarce regarding models that allow unsupervised feature extraction from brain MRI, with rare applications in cancer or psychiatric disorders(*37,38*). Therefore, radiomics – first coined by Lambin et al. as an innovative algorithm allowing the high-throughput extraction of medical images(*39*) – has remained the best way to extract features from radiological images thus far. The radiomics workflow includes a segmentation step followed by the extraction of multiple quantitative features consisting in intensity distribution, spatial relationships between intensity levels, texture patterns and shape descriptors. The most informative of these features are automatically selected from specific criteria, and can further be used in downstream tasks. The robustness of radiomics makes it applicable to multiple imaging modalities in a broad range of applications, particularly in cancer(40–42). In contrast to radiomics, which extract quantitatively measurable features, deep learning models extract more abstract and higher-level features that are expected to generate more comprehensive embeddings. Indeed, in the current study, the embeddings generated by our model generated better predictions than that based on radiomics. We explored several strategies to enhance the quality of our embeddings through data augmentation.

Our initial attempt to combine autoencoder-generated embeddings with radiomics features showed no improvement in downstream clinical predictions compared to either approach alone (data not shown). However, we discovered that incorporating synthetic MRI sequences, generated through deep learning, significantly enhanced embedding quality. This finding is particularly relevant in the current context of medical imaging, where there is increasing pressure to optimize MRI protocols by reducing acquisition times and sequences. Our results suggest that synthetic imaging could help maintain comprehensive analyses while limiting the actual scanning time. Moreover, this success with synthetic MRI data opens promising perspectives in the broader context of generative AI. Similar approaches could be developed to generate synthetic data across other modalities such as transcriptomics or pathology images (43–46,17). Such synthetic data generation could provide a powerful strategy to enhance the quality of medical imaging embeddings by incorporating complementary information from multiple modalities, even when the original data is not available.

The potential applications of *DUNE* extend to the emerging field of data fusion, where features from multiple data sources are combined to enhance prediction accuracy. Recent studies have demonstrated the power of this approach, showing how the integration of pathology and transcriptomics data can improve brain cancer prognosis prediction beyond what either modality can achieve alone (*47*). In this context, *DUNE* could provide a robust framework for incorporating radiological data into such multi-modal analyses, potentially further improving predictive performance.

While our results demonstrate *DUNE’s* broad utility, several limitations should be acknowledged. Currently, the model is optimized for three specific MRI sequences: T1, T1Gd, and FLAIR. Although additional sequences, such as diffusion-weighted imaging, could be incorporated given sufficient training data, this represents a current constraint on the model’s applicability. Furthermore, unlike the more versatile radiomics approach which can analyze any radiological image, DUNE is specifically designed for brain MRI analysis. However, the underlying architecture is fundamentally capable of processing any 3D medical image, suggesting that organ-specific versions could be developed for other clinical applications. In conclusion, *DUNE* represents a significant advance in medical image analysis, capable of extracting comprehensive low-dimensional representations from complex brain MRI scans. These embeddings effectively capture both obvious and subtle imaging features while maintaining clinical relevance. By facilitating more accurate diagnoses, refined prognostic assessments, and better-informed therapeutic decisions, *DUNE* contributes to the advancement of precision medicine in neurology. The model’s ability to generate meaningful embeddings from standard clinical imaging sequences, combined with its potential for integration into multi-modal analysis frameworks, positions it as a valuable tool for developing more personalized and effective patient care strategies.

## MATERIALS AND METHODS

### Preprocessing

Clinical and imaging data (DICOM and NIFTI files) were collected from publicly available datasets: the UK Biobank (https://www.ukbiobank.ac.uk) healthy volunteer dataset(*48*), the MCIC and COBRE schizophrenia datasets(*49,50*) (available on the SchizConnect database website: http://schizconnect.org), and the ADNI Alzheimer dataset. Glioma datasets were downloaded from The Cancer Imaging Archive (https://www.cancerimagingarchive.net)(51): UCSF-PDGM(*52*), UPenn-GBM(*53*), TCGA-LGG and TCGA-GBM(*54*).

Regarding the UKB dataset, clinical variables from the whole dataset (n≈500,000) were collected, excluding those with a missing rate ≥15%. The data were standardized and data imputation was performed using the k-Nearest Neighbors algorithm (k=140).

All the images were standardized through the preprocessing pipeline using the Advanced Normalization Tools (ANTs, https://github.com/ANTsX/ANTs) depicted in Supplemental Figure S1(*55*). When needed, DICOM files were converted to NIFTI files. Images of each sequence (T1, T1Gd, FLAIR) then underwent a series of transformations. Skull stripping was performed with the ANTS Brain extractor tool using templates and probability masks from the OASIS (Open Access Series of Imaging Studies) datasets(*56*). The N4 bias field correction algorithm was applied and FLAIR images were co-registered to T1. All images were then warped into a common space using the MNI-152 T1 1mm template (“Montreal Neurologic Institute [MNI] space”)(*57*). Pixel intensities were finally Z-score normalized. The final dimensions (X, Y, Z) of the pre-processed images were [182, 218, 160] pixels (1px/mm, 1 grayscale channel).

### Autoencoders architecture

All deep learning models were implemented using the PyTorch library (version 2.0.0). The autoencoder architectures, with the exception of the VAE model, were based on the UNet framework adapted for 3D image processing. The encoder and decoder components were built using convolutional blocks, where each block repeated twice the following sequence: a 3D convolution layer (kernel=3, stride=1, padding=1), followed by batch normalization and ReLU activation (Fig 1c). Between these convolutional blocks, max pooling layers were used in the encoder while transposed convolution layers (kernel=2, stride=2, padding=0) were used in the decoder. As the input images progressed through the encoder, their dimensions were progressively reduced while the number of channels expanded in consecutive blocks (4, 8, 16, 32, 64 and 128). The bottleneck architecture differed between conventional and variational autoencoders: conventional autoencoders used a single convolutional block, while variational autoencoders employed two separate convolutional blocks followed by a reparameterization step. Each autoencoder generated two outputs: a low-dimensional representation at the bottleneck and a reconstructed version of the input image. We set the number of encoder/decoder blocks to 6, which yielded between 1,000 and 5,000 features per input image for conventional autoencoders, and exactly 2,048 features for variational autoencoders - dimensions suitable for subsequent machine learning tasks.

### Autoencoder training algorithm

The images used for training the autoencoders were divided into train and validation sets (80%/20%). The training was carried out with a batch size of 14 images. For each batch, we aimed for the autoencoders to encode and reconstruct images as close to the original as possible. Therefore, we opted for a loss function relying on the structural similarity index measure between the input (𝑥) and the output (𝑦) produced by the autoencoders(*58*) defined as follows (1) :

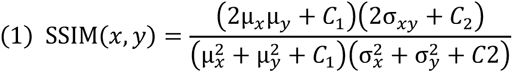

with (µ*_x_*, σ*_x_*^2^) and -µ*_y_*, σ*_y_*^2^ the respective pixel sample mean and variance of the input (𝑥) and output (𝑦) images, σ*_xy_* the covariance of 𝑥 and 𝑦, and C_1_ and 𝐶_2_ two variables to stabilize divisions with weak denominators. As the SSIM ranges from 0 to 1, the autoencoders objective is thus to minimize the following loss function (2):

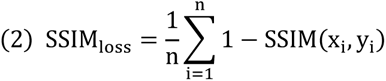

For variational autoencoders, the Kullback–Leibler divergence (KLD) loss (3) of the embedding distribution – that quantifies its divergence with the gaussian distribution – was added to the SSIM loss (4), with a weighting β parameter set to 10^-4^ (*59,60*). The ADAM algorithm was used as the optimizer, with a learning rate of 1e^-4^.

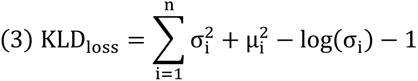

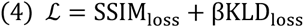

### Feature extraction

Feature extraction was performed as follows: for each input MRI sequence (T1, T1Gd, or FLAIR), features were extracted from the autoencoder’s bottleneck layer. These sequence-specific features were then concatenated to generate comprehensive patient-level embeddings. To benchmark our approach, we also extracted radiomic features (excluding diagnostic features) from each sequence using the *PyRadiomics library* (v3.1.0) (24), with whole brain masks for the segmentation step. These radiomic features (n=1,132 per sequence) were concatenated at the patient level following the same methodology as the autoencoder features, enabling direct comparison between both feature extraction methods.

### UMAP, CCA and machine learning models

Analyses were carried out with the scikit-learn library. UMAP analyses were computed on embeddings (2 components, 15 neighbors). Canonical correlations analysis were performed between embeddings and clinical variables, which were summarized to 2 main components.

Machine learning models were trained to predict clinical variables using the different generated embeddings. The model architecture was selected based on the type of clinical variable: Ridge regression for quantitative variables, L2-penalized logistic regression for categorical variables, and random survival forests for survival variables. For each clinical variable, separate models were trained using embeddings from each autoencoder architecture and radiomics features. We employed a 5-fold cross-validation procedure, with oversampling to address class imbalance in categorical variables. Hyperparameter optimization was performed using GridSearchCV within each fold to ensure optimal model configurations for each type of embedding. The predictions from all validation folds were concatenated to generate ROC curves and compute performance metrics: weighted F1-scores for categorical variables, R2 scores for quantitative variables, and C-indexes and Integrated Brier Scores for survival variables. The predictive performance of models using different embeddings were compared using Wilcoxon signed-rank tests with Bonferroni correction for multiple comparisons.

## Data availability

The source codes of the different autoencoders presented in this manuscript are available on GitHub at https://github.com/gevaertlab/BrainMR_AE_datafusion.

## ACKNOWLEDGEMENTS

This research has been conducted using the UK Biobank Resource under Application Number 51998. The results shown here are in whole or part based upon data generated by the TCGA Research Network: http://cancergenome.nih.gov

Data was downloaded from the COllaborative Informatics and Neuroimaging Suite Data Exchange tool (COINS; http://coins.mrn.org/dx) and data collection was performed at the Mind Research Network (www.mrn.org) and funded by a Center of Biomedical Research Excellence (COBRE) grant 5P20RR021938/P20GM103472 from the NIH to Dr. Vince Calhoun. The MCIC project was supported by the Department of Energy under Award Number DE-FG02-08ER64581. MCIC is the result of efforts of co-investigators from University of Iowa, University of Minnesota, University of New Mexico, Massachusetts General Hospital.

Data collection and sharing for this project were funded by the Alzheimer’s Disease Neuroimaging Initiative (ADNI) (National Institutes of Health Grant U01 AG024904) and DOD ADNI (Department of Defense award number W81XWH-12-2-0012). ADNI is funded by the National Institute on Aging, the National Institute of Biomedical Imaging and Bioengineering, and through generous contributions from the following: AbbVie, Alzheimer’s Association; Alzheimer’s Drug Discovery Foundation; Araclon Biotech; BioClinica, Inc.; Biogen; Bristol-Myers Squibb Company; CereSpir, Inc.; Cogstate; Eisai Inc.; Elan Pharmaceuticals, Inc.; Eli Lilly and Company; EuroImmun; F. Hoffmann-La Roche Ltd. and its affiliated company Genentech, Inc.; Fujirebio; GE Healthcare; IXICO Ltd.; Janssen Alzheimer Immunotherapy Research & Development, LLC.; Johnson & Johnson Pharmaceutical Research & Development LLC.; Lumosity; Lundbeck; Merck & Co., Inc.; Meso Scale Diagnostics, LLC.; NeuroRx Research; Neurotrack Technologies; Novartis Pharmaceuticals Corporation; Pfizer Inc.; Piramal Imaging; Servier; Takeda Pharmaceutical Company; and Transition Therapeutics. The Canadian Institutes of Health Research is providing funds to support ADNI clinical sites in Canada. Private sector contributions are facilitated by the Foundation for the National Institutes of Health (http://www.fnih.org). The grantee organization is the Northern California Institute for Research and Education, and the study is coordinated by the Alzheimer’s Therapeutic Research Institute at the University of Southern California. ADNI data are disseminated by the Laboratory for Neuroimaging at the University of Southern California. Data used in the preparation of this article were obtained from the Alzheimer’s Disease Neuroimaging Initiative (ADNI) database (http://adni.loni.usc.edu). As such, the investigators within the ADNI contributed to the design and implementation of ADNI and/or provided data but did not participate in the analysis or writing of this report. A complete listing of ADNI investigators can be found at: https://adni.loni.usc.edu/wp-content/uploads/how_to_apply/ADNI_Acknowledgement_List.pdf.

## AUTHOR CONTRIBUTIONS

Conceptualization: OG, TB

Methodology: OG, TB, SS, BB, MI

Investigation: TB, MI

Visualization: TB

Funding acquisition: OG

Project administration: OG

Supervision: OG

Writing – original draft: TB

Writing – review & editing: OG, FP, CS, SS

## SUPPLEMENTARY MATERIALS

**Supplemental Figure S1.**
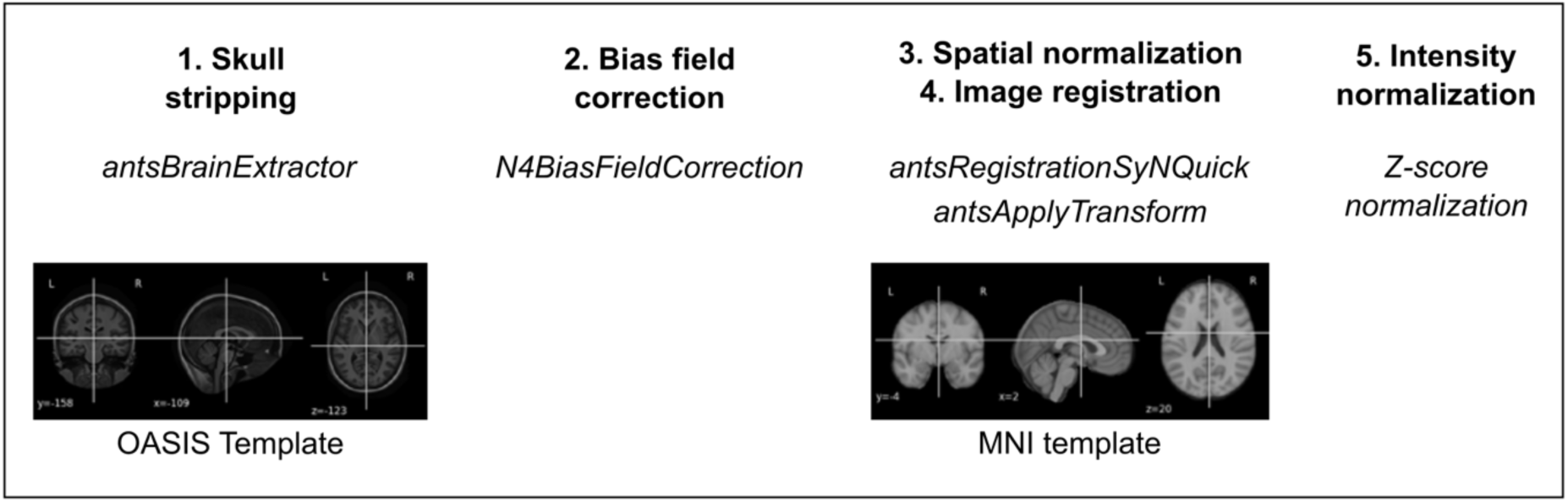
Brain MR preprocessing steps.

**Supplemental Figure S2.**
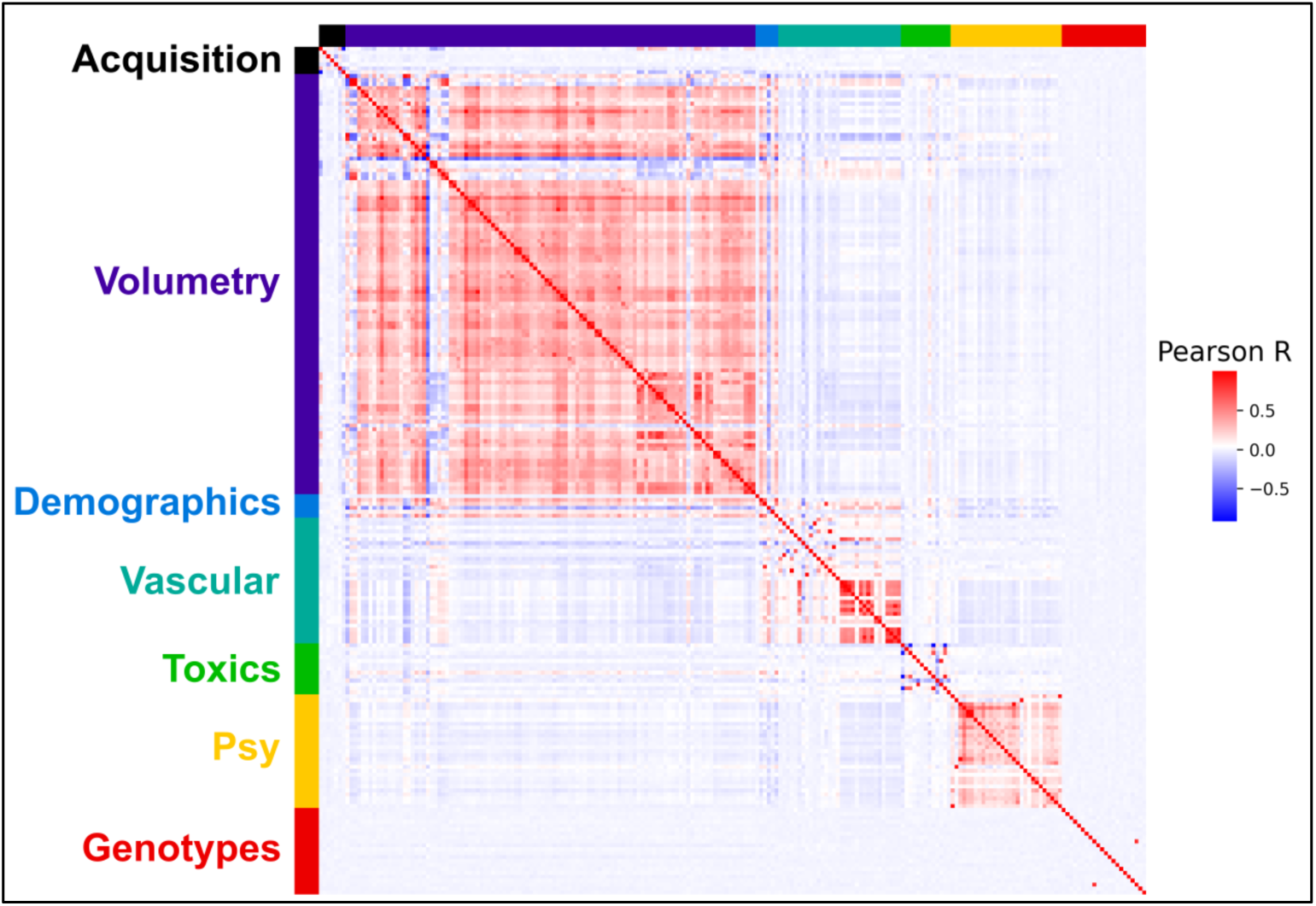
Correlation matrix of UKB clinical variables.

**Supplemental Figure S3.**
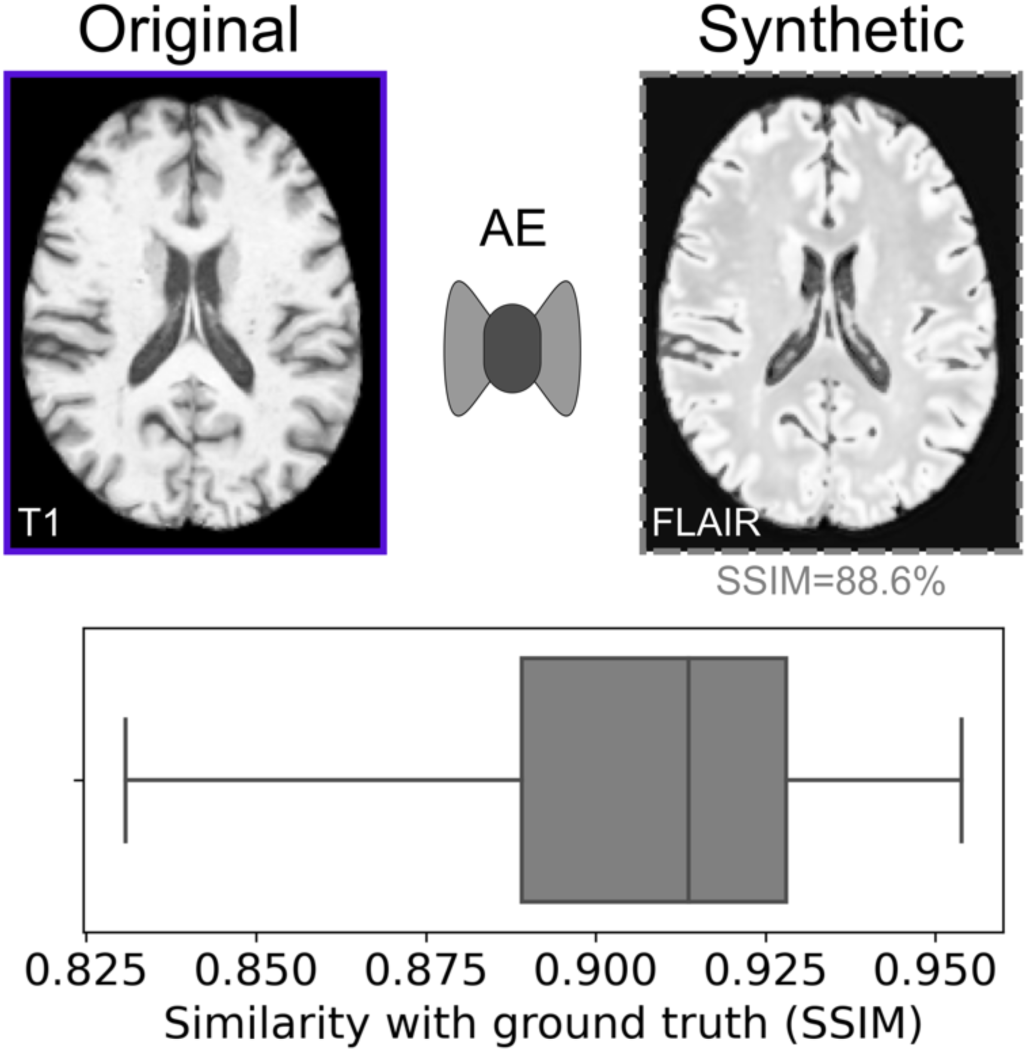
Generation of synthetic FLAIR from T1.

**Supplemental Table S1.** Brain MR datasets used in the study. Images of the UKB (n=800 cases), UPENN and UCSF datasets were used to train the models. HV: healthy volunteers; QA: quality assessment. T1Gd: T1 + gadolinium.

| Name | Sources/datasets | Normal or disease type | Sequences | Number of cases | Use |
| --- | --- | --- | --- | --- | --- |
| UKB | UK Biobank | General population | T1, FAIR | 19,955 | AE training & QA |
| UPENN | University of Pennsylvania glioblastoma dataset | Glioma (GBM only) | T1Gd, FLAIR | 612 | AE training & QA |
| UCSF | UCSF preoperative diffuse glioma MRI dataset | Glioma | T1Gd, FLAIR | 495 | AE training & QA |
| TCGA | TCGA-LGG and TCGA GBM datasets | Glioma | T1Gd, FLAIR | 168 | QA |
| ADNI | Alzheimer's Disease Neuroimaging Initiative 1 dataset | Alzheimer and HV | T1 | 818 | QA |
| SchizConnect | SchizConnect database (COBRE and MCIC datasets) | Schizophrenia and HV | T1 | 336 | QA |

